# Rapid inactivation of SARS-CoV-2 with LED irradiation of visible spectrum wavelenghts

**DOI:** 10.1101/2020.06.18.20134577

**Authors:** R. De Santis, V. Luca, G. Faggioni, S. Fillo, P. Stefanelli, G. Rezza, F. Lista

## Abstract

The COVID-19 pandemic has caused a global health crisis. The difficulty to control the viral spread due to the absence of vaccines and prophylactics measures has raised concerns about prevention and control of SARS-CoV-2. Therefore, it becomes more and more crucial the reduction of environmental risk factors through viral inactivation of aerosols and fomites. Photodynamic inactivation of microorganisms by light energy emitted in the visible spectrum region (VIS), not harmful for mammalian cells and safe for humans, has recently been described. A LED-device emitting a special combination of frequencies in the visible light spectrum was tested on SARS-CoV-2 infected cell surnatant dilutions in order to evaluate the antiviral efficacy. This preliminary *in vitro* study showed for the first time the ability to inactivate SARS-CoV-2 through LED irradiation of visible spectrum wavelengths. If further and more extensive studies will confirm these data, the usage of this LED could potentially have a big impact on the sanitization of virtually all human environments.

## Introduction

Coronavirus disease 2019 (COVID-19), a severe respiratory illness caused by a novel betacoronavirus, SARS-CoV-2 (severe acute respiratory syndrome coronavirus 2), represents currently the major global health crisis. In fact, as of middle June 2020, the COVID-19 pandemic has resulted in over 7 million cases and 413 372 deaths worldwide [WHO, 2020.]. The presence of genetic viral material on surfaces for up hours and the persistence on different materials up days in specific environmental conditions [van Doremalen et al., 2020] should suggest a significant role in the viral transmission of SARS-CoV-2 of fomites and contaminated surfaces [Sohrabi et al., 2020; Guo et al., 2020]. However, the environmental survival of the virus depends on multiple factors such as temperature, humidity, adhesion viral matrix that should be further investigated [Ratnesar-Shumate et al., 2020]. The development of systems of environmental sanitization may play a crucial role in the preventing contact infection, which represents one of the major transmission routes. Evidences of the capability of light frequencies to inactivate microorganisms have been documented by several studies, although focused on ultraviolet (UV) wavelengths. The most well-known and widespread antimicrobial irradiation model uses wavelengths within the UV spectrum, particularly in the range of 240 to 260 nm (UV-C) and has traditionally been used for surface disinfection [Blatchley, 1991]. The effectiveness of ultraviolet germicidal irradiation (UVGI), based primarily on UV-C for the inactivating a wide range of aerosolized microorganisms has been demonstrated [Nardell et al., 2008].

Furthermore, a deep ultraviolet light-emitting diode (DUV-LED) instrument generating around 250–300 nm wavelength has been reported to effectively inactivate influenza A viruses [Nishisaka-Nonaka et al., 2018] and UVB levels, representative of natural sunlight, rapidly inactivate SARS-CoV-2 on surfaces [Ratnesar-Shumate et al., 2020]. The UV lights represent however a form of radiation raising concerns of potential injury to human beings, specifically to eyes (photokeratoconjunctivitis) and skin (photodermatitis), requiring often the implementation of safety measures.

Photodynamic inactivation of microorganisms with the use of light energy emitted in the visible spectrum region (VIS) has recently been validated as a powerful means of contrasting the development of multi-drug resistant bacterial species [St Denis et al., 2011]. Evidences of the microbicidal effect of white light on bacterial strains (*H*.*pylori, P*.*mirabilis, P*.*aeruginosa*) when directly irradiated at considerable power densities (180J/cm2) and in combination with photosensitizing compounds (methylene blue) have been confirmed [Choi et al., 2010;Abu Sayem, 2014]. The use of specific frequency peaks in the blue-violet band (400-420nm) without photosensitizing factors shows microbicidal effect at lower values (36J/cm2) on a wide range of bacterial species [Maclean et al., 2009]. In the present study a device based on a combination of frequencies of the visible light was evaluated for the influence on the persistence of SARS-CoV-2.

## Methods

### Cells

Vero cells E6 were cultured at 37°C and under 5% CO2 in complete growth medium (gMEM) consisting of minimum essential medium (Life Technologies) supplemented with 10% v/v heat-inactivated fetal bovine serum (Sigma Aldrich), 2 mM Glutamax (Life Technologies), 1 mM sodium pyruvate (Life Technologies), and 1% v/v antibiotic solution (Life Technologies).

### Virus

SARS-CoV-2 was passaged once in Vero cells to generate a virus master stock. Virus master stock was then used to generate a virus working stock that was used for the experiments in the present study. The virus was propagated in Vero cells cultured in minimum essential medium (MEM) containing 2% fetal bovine serum (FBS). At 72 h after infection, virus stocks were collected by centrifuging the culture supernatants of infected Vero cells at 600 g for 5 min. The concentration of infectious virus was determined by plaque-forming titre assay. The clarified supernatants were kept at -80 °C until use.

### Exposure system

The experiments were performed using a LED strip supplied by Nextsense Srl and powered with Biovitae^®^ technology. The LED-device tested uses a special combination of frequencies in the visible light spectrum with a main peak at 413 nm (400-420nm, 430-460nm, 500-780nm). In order to prevent any sample heating during exposure, the lamp is set up with a heat sink for thermal management. To evaluate the antiviral efficacy of irradiation by visible light, a serial 10-folds dilutions of virus stock (200 ul) were placed in 96-wells plate and irradiated with 4,67 mW cm-2 at a work distance of 25 cm for a range of times (n=2 each for 40 and 60 min). The temperature and relative humidity inside the chamber were maintained within a narrow range for testing, specifically 20 ± 4°C and 19 ± 5%, respectively. Each irradiated viral stock was inoculated onto Vero monolayers in a 12-well plate in duplicates and in triplicates. After adsorption of virus for 1 h, cells were overlaid with MEM containing 1.5 % Tragacanth and 2% FBS (final concentration). Cells were incubated for 72 h in a CO_2_ incubator. An unirradiated virus suspension was used as a positive control. To calculate PFU, cells were washed with physiological solution, followed by staining with crystal violet solution. The antiviral effects of LED irradiations were calculated as (100-NLED /N0 ×100) where NLED is the PFU count of the LED-irradiated sample, and N0 is the PFU count of the sample without LED irradiation. All experiments were performed in a BSL-3 laboratory.

## Results and Discussion

The plaque assay (Figure 1) revealed that LED irradiation inactivated SARS-CoV-2. The reduction rate of the infectious titer of 97.3% was already recognized with the irradiation of the virus at 2 * 10^3^ pfu/ml for 40 minutes. When 60 minutes irradiation was used, the reduction rate was 99.8% even at the highest viral concentration 2 * 10^5^ pfu/ml (Table 1).

**Table 1.** The antiviral effects of LED irradiations at 60 and 40 minutes. The percentages of viral inactivation were calculated as (100-NLED /N0 ×100), where NLED is the PFU count of the LED-irradiated samples, and N0 is the PFU count of the sample without LED irradiation. ND not determined

| Viral titration (pfu/ml) | Time of treatment (min) |  |
| --- | --- | --- |
|  | 60 | 40 |
| $2 \times 10^5$ | 99,8 | ND |
| $2 \times 10^4$ | 99,8 | ND |
| $2 \times 10^3$ | 99,8 | 97,3 |
| $2 \times 10^2$ | 100 | 97,8 |
| $2 \times 10^1$ | 100 | 100,0 |

**Figure 1.**
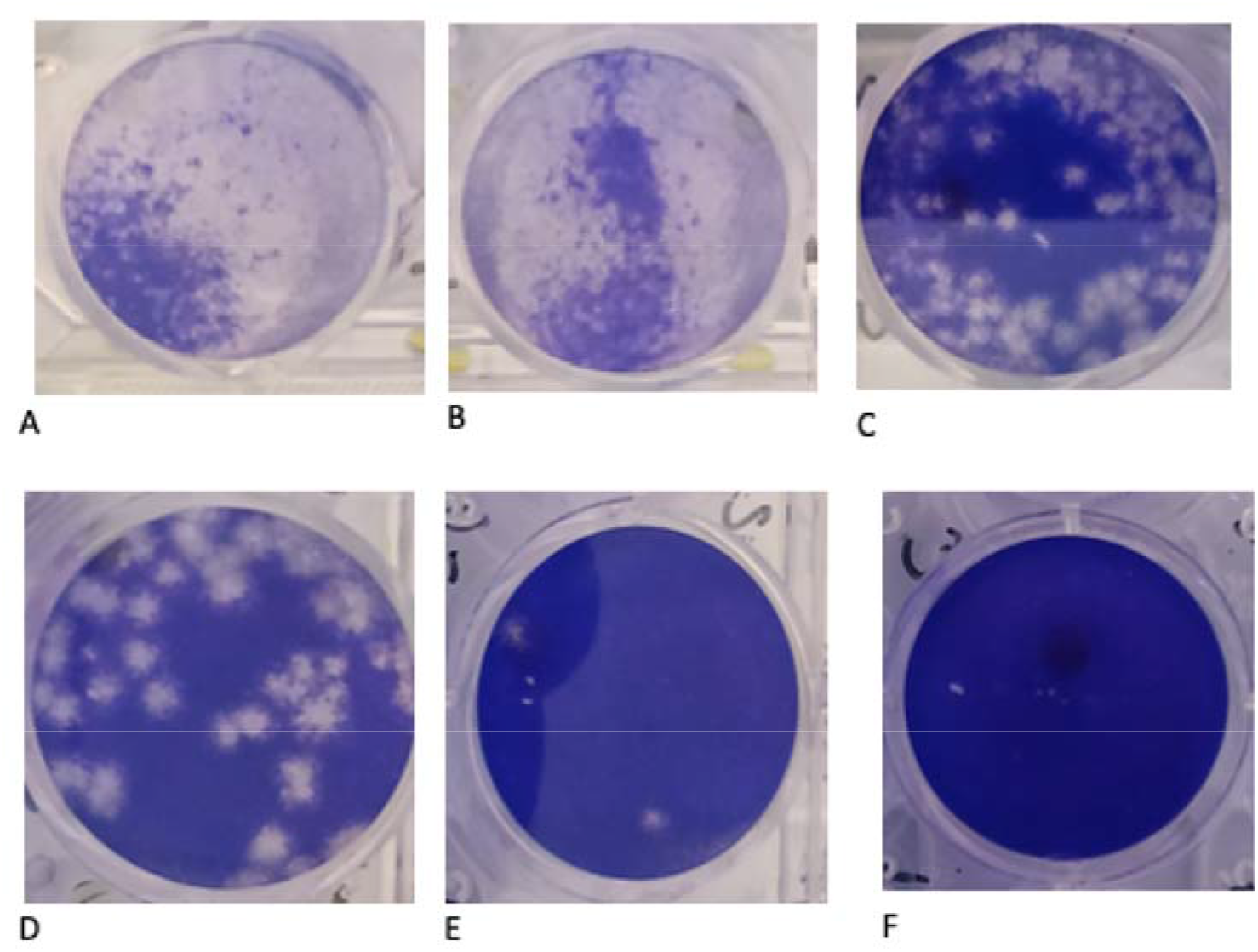
Inhibitory effects of visible light spectrum LED-irradiation on SARS-CoV-2. A-C Positive control. Plaque formation in Vero cells. (A) 10^5^ pfu/ml; (B) 10^4^ pfu/ml; (C) 10^3^ pfu/ml. D-E. The viral dilutions irradiated with LED for 60 min and inoculated to Vero cells. (D) 2x 10^2^ pfu/ml; (E) 10 pfu/ml; (F) 0 pfu/ml. As showed in D, the infectious titer reduction rate was 99.8%.

The photodynamic inactivation of microorganisms with the use of light energy emitted in the region of the visible spectrum has long been known. The wavelengths emitted by a light energy in some monochromatic peaks of the visible spectrum, cause the excitation of photo-sensitizing molecules (porphyrins), that determine the production of singlet oxygen and other reactive species (RS) which, reacting with intracellular components, damaging DNA and the plasma membrane. The LED-device tested providing combination of frequencies of the visible light spectrum (400-420nm, 430-460nm, 500-780nm), can inactivate a wide range of bacterial species GRAM + (*S*.*aureus, MRSA, S*.*epidermidis, S*.*pyogenes, E*.*faecalis, C*.*perfringens*) and GRAM– (*A*.*baumannii, P*.*aeruginosa, E*.*coli, P*.*vulgaris, K*.*pneumoniae*) [Maclean et al., 2009]. Furthermore, the exposition of the mammalian cells to bactericidal irradiance levels emitted through these low energy devices, has no significant effect on normal cell function [Thurman et al., 2019; Ramakrishnan et al., 2015]. Therefore, on the basis on these data, the possibility of developing new rapid viral inactivation method capable of reducing infections through aerosols and contact transmission, drove us to perform this study. The preliminary *in vitro* experiments demonstrated for the first time the inactivation of SARS-CoV-2 under the visible light spectrum irradiation of the LED-device. These devices, emitting a white light, may be used both for the sanitization in virtually all human environments as well as lighting devices. Further experiments in a multicentric format, using different biological matrices, as other respiratory viruses such as influenza virus, and environmental conditions will be carried out in order to confirm this new virucidal technology that could improve significantly public health.

## Data Availability

the Authors declare the data availability

## Ackowlegements

the authors thank Elisa Regalbuto, Nino D’Amore, Filippo Molinari, Giancarlo Petralito for for the virus SarsCoV2 titration and technical assistance, Stefano Fiore, Eleonora Benedetti, Concetta Fabiani, Antonella Marchi, for the virus SarsCoV2 cultivation technical assistance.

## References

Blatchley, E. R., III, and M. M. Peel. 1991. “Disinfection by ultraviolet irradiation.” p. 823–851. In S. S. Block (ed.), Disinfection, sterilisation and preservation, 4th ed. Lea and Febiger, Philadelphia, PA

Choi SS, Lee HK, Chae HS. “In vitro photodynamic antimicrobial activity of methylene blue and endoscopic white light against Helicobacter pylori 26695.” J Photochem Photobiol B. 2010 Dec 2;101(3):206–9. doi:10.1016/j.jphotobiol. 2010.07.004. Epub 2010 Jul 15.

Guo ZD, Wang ZY, Zhang SF, et al. Aerosol and surface distribution of severe acute respiratory syndrome corona-virus 2 in hospital wards, Wuhan, China, 2020 [published online ahead of print 10 April 2020]. Emerg Infect Dis doi: 10.3201/eid2607.200885.

Michelle Maclean, Scott J. MacGregor, John G. Anderson and Gerry Woolsey “Inactivation of Bacterial Pathogens following Exposure to Light from a 405-Nanometer Light-Emitting Diode Array” Appl. Environ. Microbiol. April 2009 vol. 75 no. 7 1932–1937. doi:10.1128/AEM.01892-08 Nardell EA, Bucher SJ, Brickner PW, Wang C, Vincent RL, Becan-McBride K, James MA, Michael M, Wright JD. Safety of Upper-Room Ultraviolet Germicidal Air Disinfection for Room Occupants: Results from the Tuberculosis Ultraviolet Shelter Study. Public Health Rep. 2008 Jan-Feb;123(1):52–60. doi: 10.1177/003335490812300108.

Nishisaka-Nonaka R, Mawatari K, Yamamoto T, et al. Irradiation by ultraviolet light-emitting diodes inactivates influenza a viruses by inhibiting replication and transcription of viral RNA in host cells. J Photochem Photobiol B. Biology 2018 Dec;189:193–200

Ramakrishnan P, Maclean M, MacGregor SJ, Anderson JG, Grant MH. Cytotoxic Responses to 405nm Light Exposure in Mammalian and Bacterial Cells: Involvement of Reactive Oxygen Species. Toxicol In Vitro. 2016 Jun;33:54–62. doi: 10.1016/j.tiv.2016.02.011.

Sohrabi C, Alsafi Z, O’Neill N, et al. World Health Organization declares global emergency: A review of the 2019 novel coronavirus (COVID-19). Int J Surg 2020; 76:71–6

St Denis TG, Dai T, Izikson L, Astrakas C, Anderson RR, Hamblin MR, Tegos GP. “All you need is light: antimicrobial photoinactivation as an evolving and emerging discovery strategy against infectious disease.” Virulence. 2011 Nov-Dec;2(6):509–20. doi: 10.4161/viru.2.6.17889. Epub 2011 Nov 1. DOI:10.4161/viru.2.6.17889

Shanna Ratnesar-Shumate, Gregory Williams, Brian Green, Melissa Krause, Brian Holland, Stewart Wood, Jordan Bohannon, Jeremy Boydston, Denise Freeburger, Idris Hooper, Katie Beck, John Yeager, Louis A. Altamura, Jennifer Biryukov, Jason Yolitz, Michael Schuit, Victoria Wahl, Michael Hevey, and Paul Dabisch. Simulated Sunlight Rapidly Inactivates SARS-CoV-2 on Surfaces. J Infect Dis. 2020 May 20;jiaa274. doi: 10.1093/infdis/jiaa274. Online ahead of print.

Thurman CE, Muthuswamy A, Klinger MM, Roble GS. Safety Evaluation of a 405-nm LED Device for Direct Antimicrobial Treatment of the Murine Brain. Comp Med. 2019 Aug 1;69(4):283–290. doi: 10.30802/AALAS-CM-18-000126

van Doremalen N, Bushmaker T, Morris DH, et al. Aerosol and surface stability of SARS-CoV-2 as compared with SARS-CoV-1. N Engl J Med 2020; 382:1564–7.

